# Long COVID in a highly vaccinated population infected during a SARS-CoV-2 Omicron wave – Australia, 2022

**DOI:** 10.1101/2023.08.06.23293706

**Authors:** Mulu Woldegiorgis, Gemma Cadby, Sera Ngeh, Rosemary Korda, Paul Armstrong, Jelena Maticevic, Paul Knight, Andrew Jardine, Lauren Bloomfield, Paul Effler

## Abstract

**Objective:** To characterise Long COVID in a highly vaccinated population infected by Omicron.

**Design:** Follow-up survey of persons testing positive for SARS-CoV-2 in Western Australia, 16 July-3 August 2022.

**Setting:** Community

**Participants:** 22,744 persons with COVID-19 who had agreed to participate in research at the time of diagnosis were texted a survey link 90 days later; non-responders were telephoned. Post stratification weights were applied to responses from 11,697 (51.4%) participants, 94.0% of whom had received >= 3 vaccine doses.

**Main outcome measures:** Prevalence of ‘Long COVID’ - defined as reporting new or ongoing COVID-19 illness-related symptoms or health issues 90 days post diagnosis; associated health care utilisation, reductions in work/study and risk factors were assessed using log-binomial regression.

**Results:** 18.2% (n=2,130) of respondents met case definition for Long COVID. Female sex, being 50-69 years of age, pre-existing health issues, residing in a rural or remote area, and receiving fewer vaccine doses were significant independent predictors of Long COVID (p < 0.05). Persons with Long COVID reported a median of 6 symptoms, most commonly fatigue (70.6%) and difficulty concentrating (59.6%); 38.2% consulted a GP and 1.6% reported hospitalisation in the month prior to the survey due to ongoing symptoms. Of 1,778 respondents with Long COVID who were working/studying before their COVID-19 diagnosis, 17.9% reported reducing/discontinuing work/study.

**Conclusion:** 90 days post Omicron infection, almost 1 in 5 respondents reported Long COVID symptoms; 1 in 15 of all persons with COVID-19 sought healthcare for associated health concerns >=2 months after the acute illness.

**Significance of the study:** *The known:* The prevalence of Long COVID varies widely across studies conducted in diverse settings globally (range: 9%-81%).

*The new:* In a highly vaccinated population (94% with >=3 vaccine doses), almost 20% of persons infected with the SARS-CoV-2 Omicron variant reported symptoms consistent with Long COVID 90 days post diagnosis. Long COVID was associated with sustained negative impacts on work/study and a substantial utilisation of GP services 2-3 months after the acute illness; however, ED presentations and hospitalisations for Long COVID were rare.

*The implications:* GP clinics play a significant role in managing the burden of Long COVID in Australia.

## Introduction

The SARS-CoV-2 pandemic continues to cause significant morbidity and mortality globally (1, 2). While the most severe symptoms of COVID-19 tend to occur during the acute phase of infection, numerous studies have shown that a substantial proportion of patients continue to experience persistent symptoms for several weeks or months following the acute illness, a condition commonly referred to as “Long COVID” or “post COVID-19 condition” (3, 4). Long COVID can affect multiple organ systems, resulting in respiratory, neurological, cardiovascular, and gastrointestinal symptoms which makes precisely defining what constitutes Long COVID challenging (5). Due to differences in the number and duration of symptoms required to meet the various case definitions used for long COVID, direct comparison of results across studies has been difficult (6, 7). To help minimize such disparities, the World Health Organization (WHO) used a consensus process to develop a clinical case definition for Long COVID in 2021 (8).

Western Australia (WA) remained virtually free of locally acquired COVID-19 illnesses until late February 2022 when inter-jurisdictional and overseas travel restrictions were relaxed, by which time >90% of the vaccine-eligible population had been immunised (9). From a global perspective, WA’s experience is rather unique as virtually all COVID-19 illnesses in 2022 were caused by the Omicron variant among a population lacking any background immunity from previous infection due to earlier SARS-CoV-2variants (10, 11). Therefore, estimates of the risk for developing Long COVID derived from other settings may not be applicable to WA. The aim of this study was to describe Long COVID and its impacts among a highly vaccinated population infected exclusively by Omicron.

## Methods

### Participants

All WA residents aged 18 years and over with a COVID-19 infection diagnosed by polymerase chain reaction (PCR) or rapid antigen test (RAT) who were reported to the WA Department of Health (DOH) between 16 July and 03 August 2022 and who had consented to be contacted for research as part of the original case interview process were eligible for the study. Individuals with more than one SARS-CoV-2 infection were excluded.

The DOH sent a text message with a unique link to an online survey to all eligible persons 90 days after the onset date for their COVID-19 illness. The text messages were sent between 14 October and 01 November 2022. Persons who did not respond to the initial text message within 24 hours were sent up to two reminder messages. Trained DOH interviewers made follow-up telephone calls to those who did not respond to reminder text messages (Figure 1). Survey responses were linked to demographic and vaccination information collected as part of the initial COVID-19 disease notification and case investigation process.

**Figure 1:**
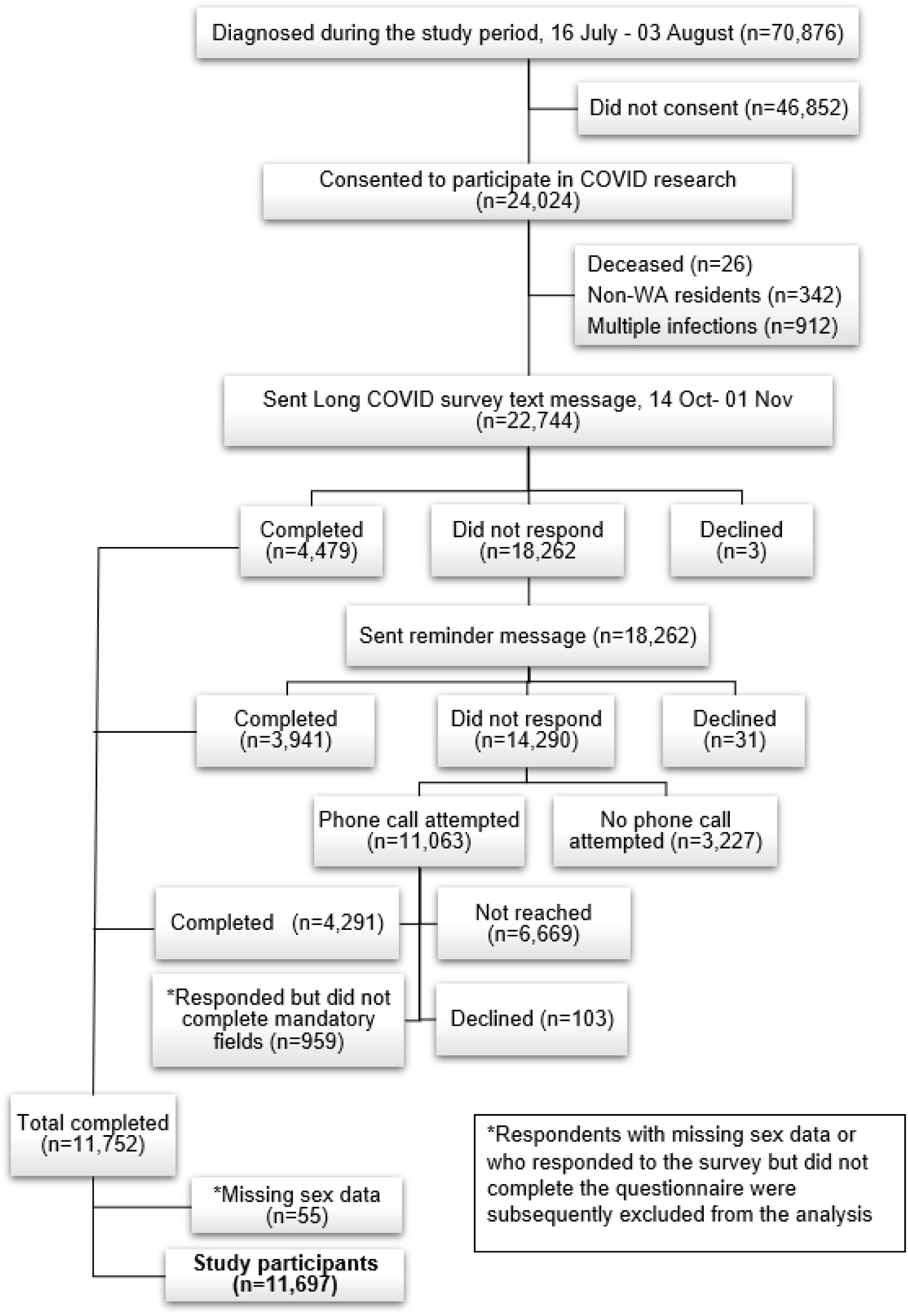
CONSORT Flow diagram: Long COVID survey participant recruitment process and response status, Western Australia, 14 October to 01 November 2022

### Outcome measures

The study had three principal outcome measures. The first was the proportion of respondents with Long COVID, defined as the presence of new or ongoing COVID-19 illness-related symptoms or health issues among persons who reported not being fully recovered 90 days post diagnosis. The second was the proportion of persons with Long COVID who reported seeking health care services for ongoing symptoms in the month before completing the survey, i.e. 2 to 3 months after an acute COVID-19 illness. The third was the proportion of respondents who were working or studying prior to their COVID-19 diagnosis who reported not fully returning to work or study due to Long COVID. In addition, the survey captured information on pre-existing health issues and specific symptoms commonly associated with Long COVID in other settings (See Appendix 1 for details).

### Exposure variables

Information collected regarding potential exposures associated with Long COVID included sex (male/female), age group (18-29/30-39/40-49/50-59/60-69/≥70 years old), pre-existing health issues prior to COVID-19 infection (yes/no), area of residence (regional-remote/metropolitan), and number of COVID-19 vaccine doses received >= 1 week prior to COVID-19 diagnosis (0-2 doses/3 doses/≥4 doses).

### Data Analysis

Sampling weights were calculated using sex and 10-year age-groups to adjust for over- or under-representation of some age/sex cohorts among the survey respondents compared to all adults diagnosed with SARS-CoV-2 during the study period, i.e., the target population. The weighted data were used to calculate proportions adjusted to the target population. Further information on the sampling weights is available in Appendix 2.

Log-binomial regression was used to examine the relationship between potential predictors of Long COVID and other outcomes of interest, while controlling for other exposure variables. We used a p-value threshold of <0.05 for determining statistical significance. Statistical analyses were performed in Stata 15. For people who did not have sex recorded in the initial case investigation database, their sex was imputed using the ‘sex’ library in Rv4.2.1, utilising first names and US Census data from 1950-2005 (12). Respondents whose sex could not be imputed and others who did not complete the questionnaire were excluded from the analysis (Figure 1).

The study was approved by the Department of Health WA Human Research Ethics Committee (PRN:RGS0000005516).

## Results

All 70,876 adults with COVID-19 reported to the DOH during the study period, were asked for permission to be contacted for future research and 24,024 individuals (33.9%) consented. Of these, 1,280 (5.3%) were excluded because they had >1 SARS-CoV-2 infection reported (n=912), were not residents of WA (n=342) or were deceased at the time of the survey (n=26). The remaining 22,744 persons were sent the SMS inviting them to participate and 12,711 (55.8%) agreed; data from 55 (0.4%) and 959 (7.5%) of those who agreed were subsequently excluded due to missing information on sex or incomplete survey responses, respectively, resulting in a final ‘study population’ of 11,697 respondents (51.4% of those invited to participate). (Figure 1). Characteristics of the cohort invited to participate compared to the study population are shown in Table 1.

**Table 1:**
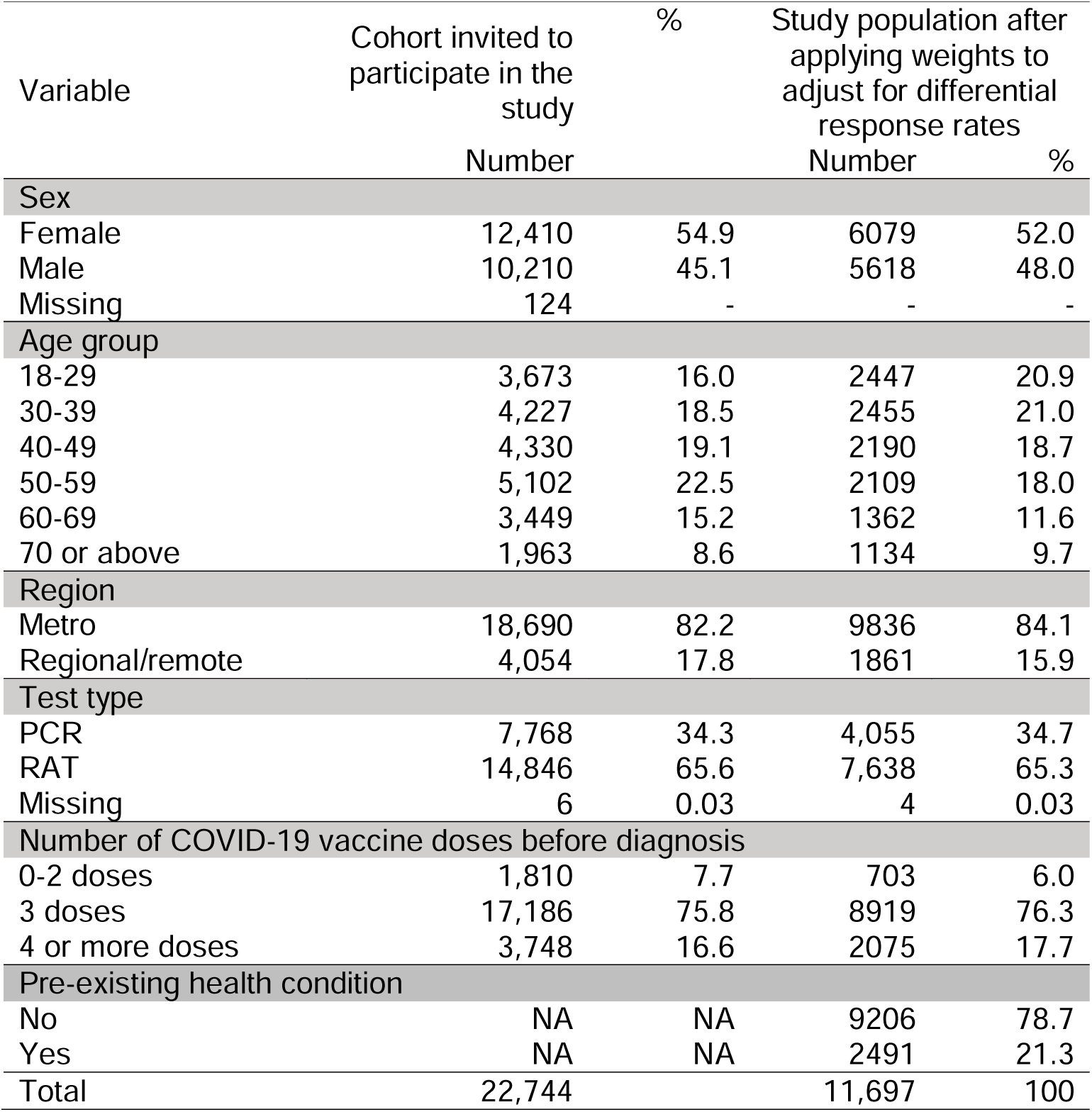
Characteristics of Long COVID survey participants in Western Australia between 14 October and 01 November 2022.

| Variable | Cohort invited to participate in the study | % | Study population after applying weights to adjust for differential response rates |  |
| --- | --- | --- | --- | --- |
|  | Number |  | Number | % |
| Sex |  |  |  |  |
| Female | 12,410 | 54.9 | 6079 | 52.0 |
| Male | 10,210 | 45.1 | 5618 | 48.0 |
| Missing | 124 | - | - | - |
| Age group |  |  |  |  |
| 18-29 | 3,673 | 16.0 | 2447 | 20.9 |
| 30-39 | 4,227 | 18.5 | 2455 | 21.0 |
| 40-49 | 4,330 | 19.1 | 2190 | 18.7 |
| 50-59 | 5,102 | 22.5 | 2109 | 18.0 |
| 60-69 | 3,449 | 15.2 | 1362 | 11.6 |
| 70 or above | 1,963 | 8.6 | 1134 | 9.7 |
| Region |  |  |  |  |
| Metro | 18,690 | 82.2 | 9836 | 84.1 |
| Regional/remote | 4,054 | 17.8 | 1861 | 15.9 |
| Test type |  |  |  |  |
| PCR | 7,768 | 34.3 | 4,055 | 34.7 |
| RAT | 14,846 | 65.6 | 7,638 | 65.3 |
| Missing | 6 | 0.03 | 4 | 0.03 |
| Number of COVID-19 vaccine doses before diagnosis |  |  |  |  |
| 0-2 doses | 1,810 | 7.7 | 703 | 6.0 |
| 3 doses | 17,186 | 75.8 | 8919 | 76.3 |
| 4 or more doses | 3,748 | 16.6 | 2075 | 17.7 |
| Pre-existing health condition |  |  |  |  |
| No | NA | NA | 9206 | 78.7 |
| Yes | NA | NA | 2491 | 21.3 |
| Total | 22,744 |  | 11,697 | 100 |

### Risk of Long COVID and associated symptoms

A total of 2,130 (18.2%, 95%CI: 17.5%-18.9%) study participants were classified as having Long COVID at 90 days post diagnosis (Table 2). After adjusting for all exposure variables in the log-binomial regression model, females had a 50% higher risk of developing Long COVID compared to males (RR=1.5, 95% CI: 1.4-1.6), and the risk increased with age, with individuals aged 50-69 years having a >= 50% higher risk compared to those aged 18-29 years (RR: 1.6, 95%CI:1.4-1.9). Individuals with pre-existing health issues also had a 50% higher risk of Long COVID compared to individuals reporting no pre-existing health issues (RR=1.6, 95% CI: 1.4-1.7) and persons residing in regional/remote areas had a 10% higher risk compared to persons residing in metropolitan Perth (RR=1.1, 95% CI: 1.0-1.2). Finally, there was a significant inverse relationship between the number of COVID-19 vaccine doses received and the risk of having Long COVID; individuals receiving 2 or fewer and 3 doses of COVID-19 vaccine were 60% (RR=1.6, 95%CI: 1.3-1.9) and 40% (RR=1.4, 95%CI: 1.3-1.6) more likely to have Long COVID compared to those receiving ≥4 doses (Table 2).

**Table 2:** Factors associated with Long COVID among survey participants in Western Australia 90 days post COVID-19 diagnosis, 14 October - 01 November 2022.

| Variable | *Number with Long COVID/total (%) | Adj. RR (95% CI) | P-value |
| --- | --- | --- | --- |
| Total | 2,130/11,697 (18.2) | - | - |
| Sex (ref= Male) |  |  |  |
| Male | 796/5,618 (14.2) | - | - |
| Female | 1,334/6,080 (21.9) | 1.5 (1.4, 1.6) | <0.001 |
| Age group (ref= 18-29) |  |  |  |
| 18-29 | 347/2,447 (14.2) | - | - |
| 30-39 | 416/2,455 (16.9) | 1.2 (1.0, 1.4) |  |
| 40-49 | 418/2,191 (19.1) | 1.3 (1.2, 1.6) |  |
| 50-59 | 460/2,109 (21.8) | 1.5 (1.3, 1.8) | <0.001 |
| 60-69 | 295/1,363 (21.6) | 1.6 (1.4, 1.9) |  |
| 70 and above | 194/1,134 (17.1) | 1.4 (1.1, 1.7) |  |
| Area of residence (ref=Metropolitan) |  |  |  |
| Metropolitan | 1,749/9,835 (17.8) | - | - |
| Regional and remote | 380/1,861 (20.4) | 1.1 (1.0, 1.2) | 0.043 |
| COVID-19 vaccination doses (ref= Four or more doses) |  |  |  |
| Four or more doses | 337/2075 (16.2) | - | - |
| Three doses | 1,652/8,919 (18.5) | 1.4 (1.3, 1.6) | <0.001 |
| Zero to two doses | 140/703 (19.9) | 1.6 (1.3,1.9) |  |
| Any pre-existing health issues (ref=No) |  |  |  |
| No | 1,493/9,206 (16.2) | - | - |
| Yes | 637/2,491 (25.6) | 1.5 (1.4, 1.7) | <0.001 |

95%CI= 95% confidence interval, RR= Relative risk, adj.=adjusted for other covariates included in the table, Ref=Reference group
\*These numbers are weighted

More than 90% of persons with Long COVID were polysymptomatic; the median number of reported symptoms was six (interquartile range [IQR]=3-9) (Figure 2). The proportion of persons reporting specific Long COVID symptoms is shown in Figure 3; a majority reported “tiredness or fatigue that interfered with daily life” (70.6%) and “difficulty thinking or concentration” (59.6%). One-third (32.6%) of female participants aged 18-49 years old reported changes in their menstrual cycle.

**Figure 2:**
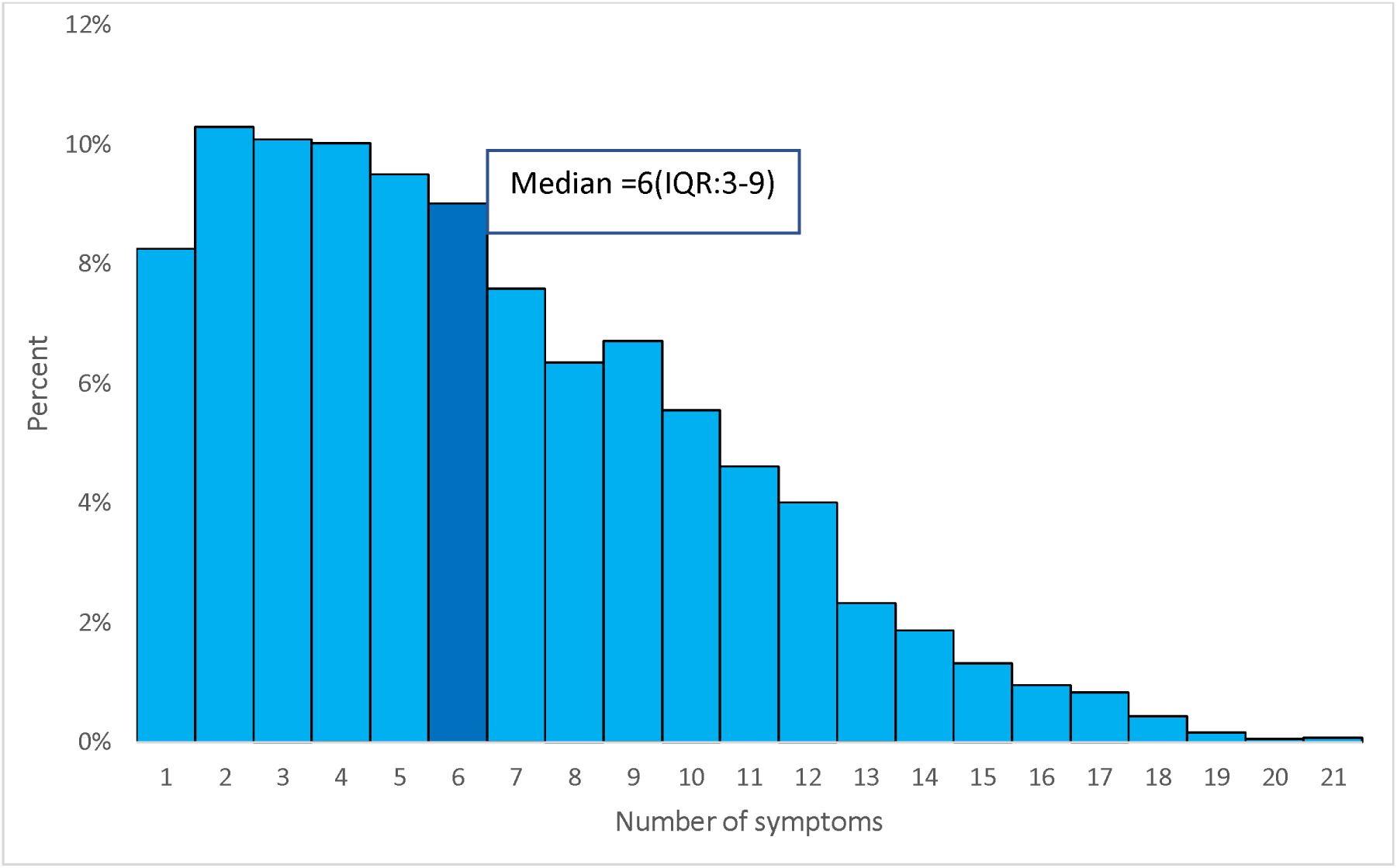
Number of symptoms reported by survey participants with Long COVID in Western Australia, 14 October – 01 November 2022

**Figure 3:**
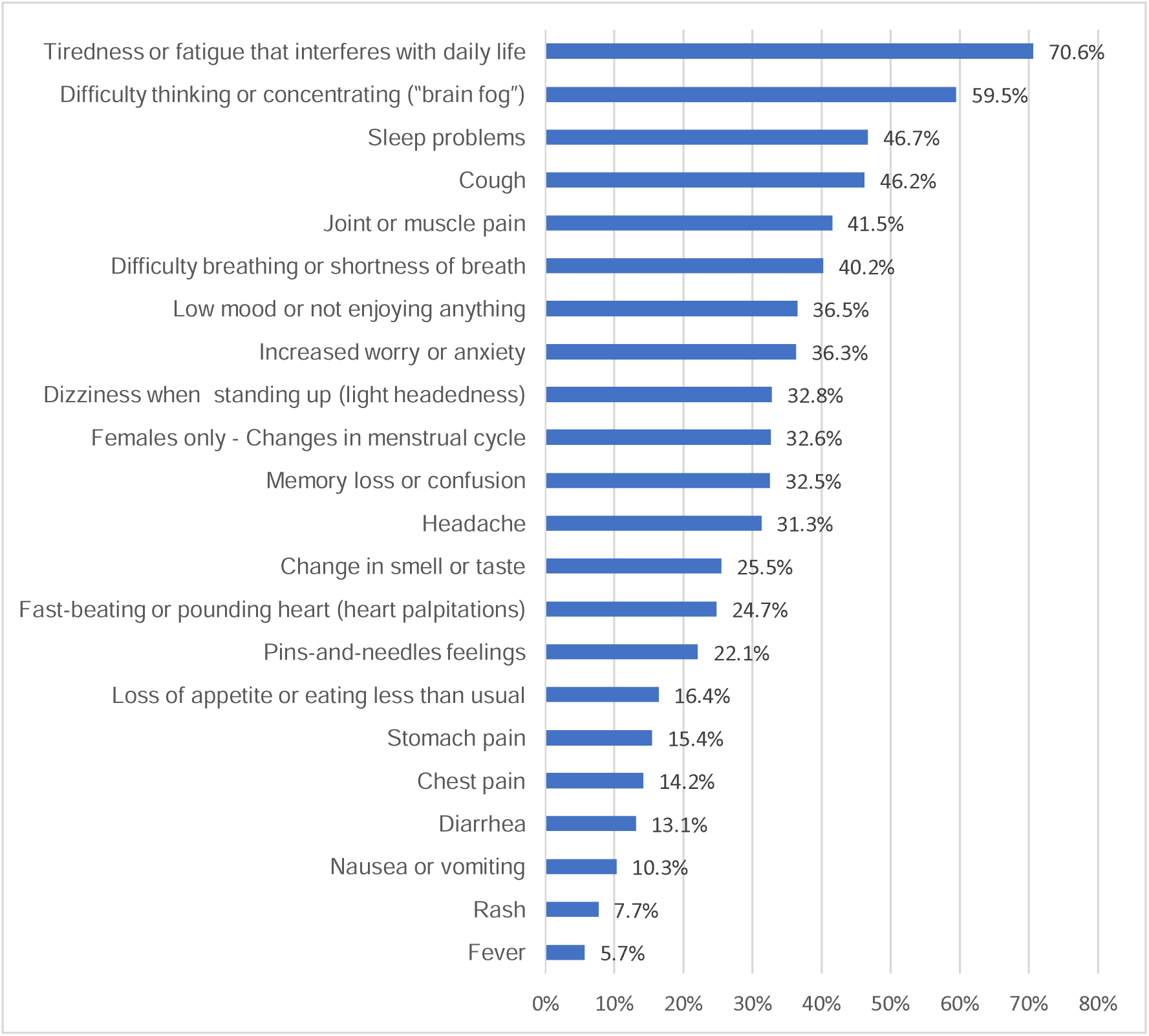
Frequency of symptoms reported by survey participants with Long COVID in Western Australia, 14 October - 01 November 2022

### Health service use among participants with long COVID

Almost 40% of people with Long COVID reported using health services in the month preceding the survey because of ongoing symptoms following their COVID-19 illness (Table 3). Of these, 38.2% visited a GP, 3.9% presented to ED, 1.6% were admitted to hospital; a few individuals reported a combination of GP visits, ED visits and/or hospital admission. After adjusting for all exposure variables, females were 20% more likely to report using health services due to ongoing COVID-related symptoms compared to males (RR=1.2, 95%CI: 1.1-1.4); no other characteristics were associated with health service utilization (Table 3).

**Table 3:**
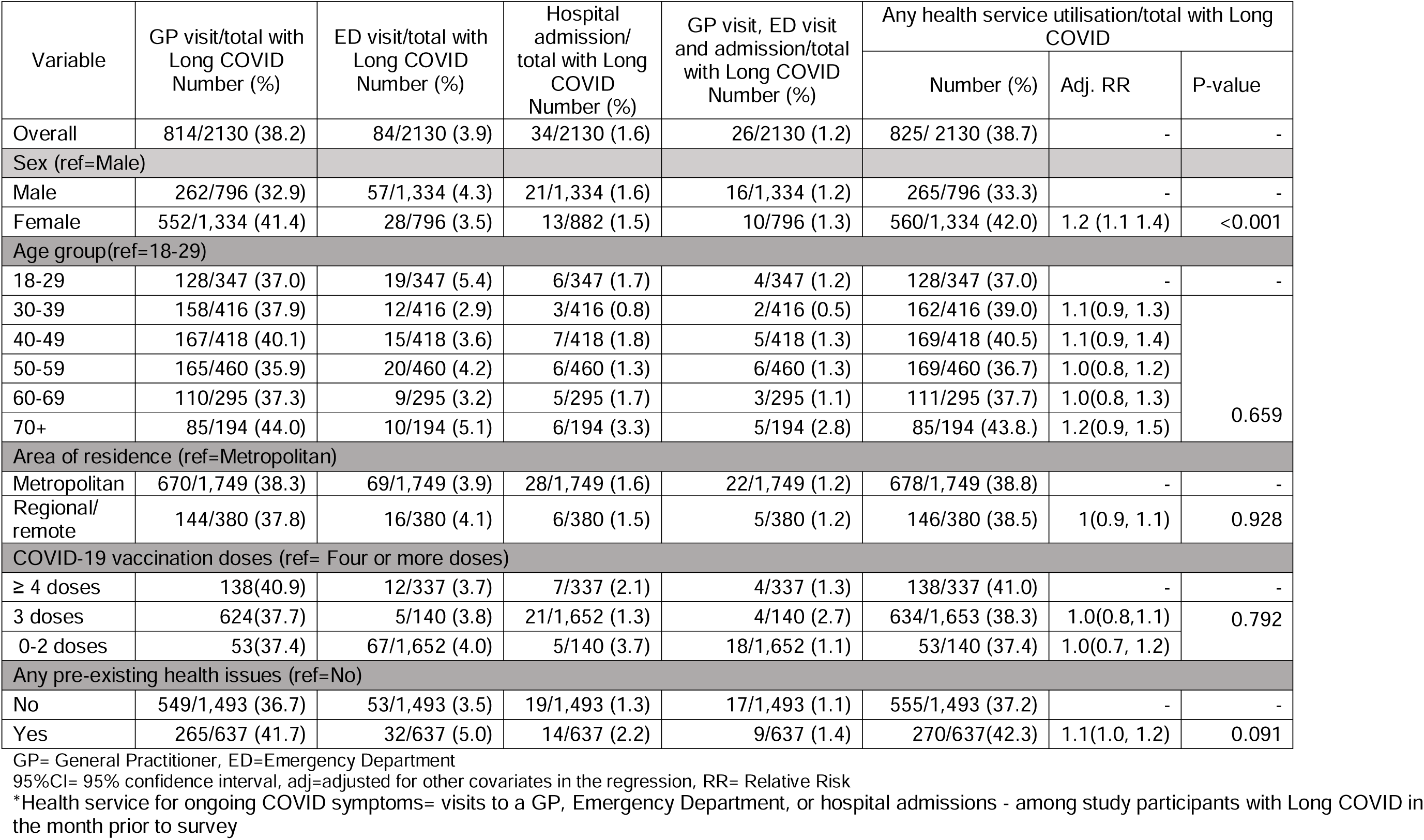
Factors associated with health service use for ongoing COVID symptoms among survey participants with Long COVID in Western Australia, 14 October – 01 November 2022.

### Self-reported health status

A majority of individuals with Long COVID reported their health status 90 days post diagnosis as “fair” (49.9%) or “poor” (9.7%); in contrast, among individuals without Long COVID, a notably greater proportion indicated their health status as excellent (23.2%) or good (58.9%) (Figure 4).

**Figure 4:**
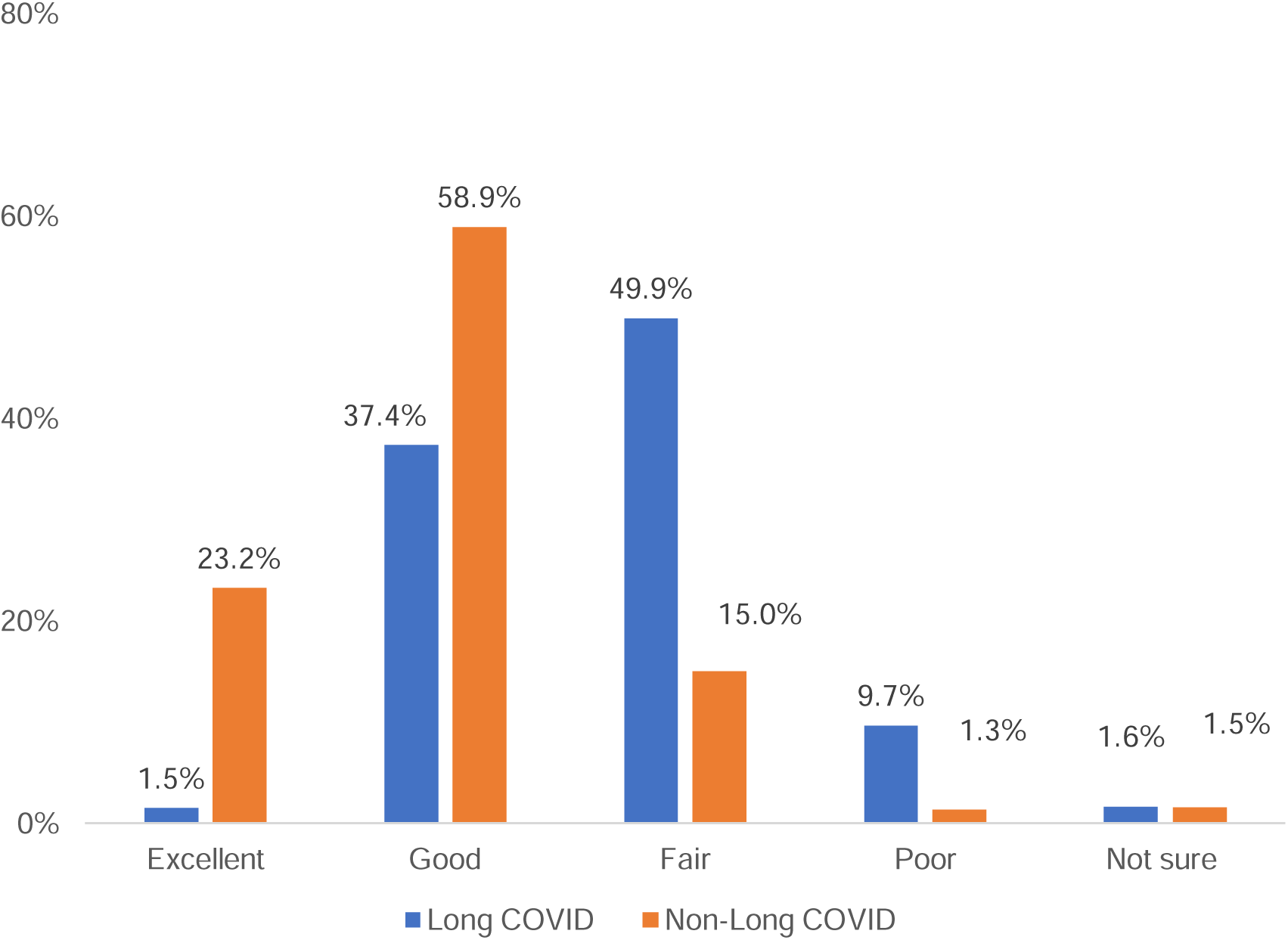
Health status at three months after COVID-19 diagnosis among survey participants with and without Long COVID

### Return to work or study

Of the 1,702 respondents with Long COVID who were working/studying before their COVID-19 diagnosis, 64.6% (1,100/1,702) returned to work/study within a month after their COVID-19 diagnosis. A further 17.2% (293/1,702) had returned to their previous work/study hours 90 days post COVID-19 diagnosis; 15.6% (265/1,702) were working/studying but had reduced their hours, and the remaining 2.6% (44) had not returned to work/study (Figure 5). After adjusting for other participant characteristics, individuals who received 0-2 doses of COVID-19 vaccine were 70% more likely to have reduced work hours or not returned to work/study 90 days post COVID-19 diagnosis compared to those who received 4 or more doses (RR=1.7, 95%CI: 1.1-2.7). Females (RR=1.3, 95%CI: 1.0-1.6) and persons with pre-existing health issues (RR=1.5, 95%CI: 1.2-1.9) were also less likely to fully return to work 90 days after their COVID-19 diagnosis (Table 4).

**Figure 5:**
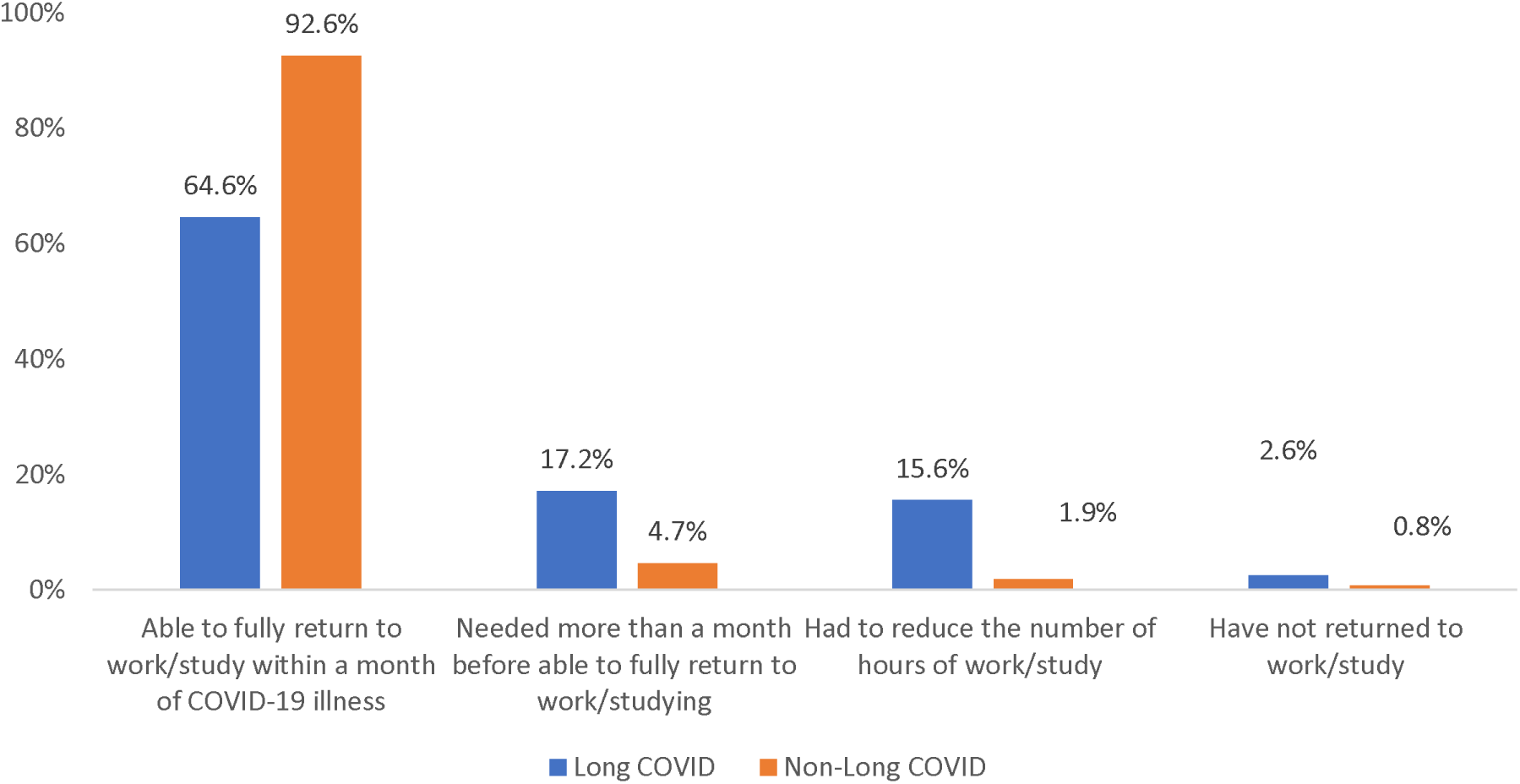
Work/study status among survey participants with and without Long COVID three months after COVID diagnosis

**Table 4:** Factors associated with non-return to study/work or reduced number of hours among survey participants with Long COVID who were working or studying prior to COVID-19 diagnosis, Western Australia, 14 October – 01 November 2022.

| Covariates | Not returned to study or<br>work or reduced hours<br>/Total with Long COVID (%) | Adj. RR (95% CI) | P-value |
| --- | --- | --- | --- |
| Total | 318/1,778(17.8) |  |  |
| Sex (ref=Male) |  |  |  |
| Male | 103/690(14.9) | - | - |
| Female | 215/1,089(19.8) | 1.3(1.0,1.6) | 0.040 |
| Age group (ref=18-29) |  |  |  |
| 18-29 | 71/333(21.2) | - | - |
| 30-39 | 63/383(16.3) | 0.8(0.5,0.1) | 0.031 |
| 40-49 | 75/393(19.0) | 1.0(0.7,1.3) |  |
| 50-59 | 61/416(14.6) | 0.7(0.5,1.0) |  |
| 60-69 | 33/200(16.4) | 0.8(0.5,1.1) |  |
| 70 and above | 16/51(31.7) | 1.5(0.9,2.5) |  |
| Area of residence (ref=Metropolitan) |  |  |  |
| Metropolitan | 259/1,461(17.7) | - | - |
| Regional and remote | 60/318(18.7) | 1.1(1.0,1.2) | 0.458 |
| COVID-19 vaccination doses (ref= Four or more doses) |  |  |  |
| Four or more doses | 38/180(21.1) | - | - |
| Three doses | 246/1,490(16.5) | 0.9(0.6,1.2) | <0.001 |
| Zero to two doses | 34/109(31.7) | 1.7(1.1,2.7) |  |
| Any pre-existing health issues (ref=No) |  |  |  |
| No | 201/1,308(15.4) | - | - |
| Yes | 117/470(24.9) | 1.5(1.2,1.9) | <0.001 |

95%CI= 95% confidence interval, RR= Relative risk, adj=adjusted for other exposure variables in the regression, ref=Reference group.

## Discussion

There are several important findings from this investigation. First, in a highly vaccinated population exposed exclusively to the Omicron variant, almost 20% reported ongoing symptoms compatible with Long COVID at 90 days post COVID-19 diagnosis. This figure is substantially higher than the prevalence reported from a review of Australian data from earlier in the pandemic which found 5% to 9.7% of persons with SARS-CoV-2 experienced ‘post COVID condition’ at 12 weeks or more after infection (13). The proportion also exceeds the those reported from large studies in the United Kingdom and Canada (14–17). The WA results are however similar to those from a recent study from Queensland, where 21% of persons diagnosed with Omicron reported ongoing symptoms at 12 weeks (18). Thus, while limited evidence from Australia and other countries has suggested that the risk of Long COVID may be lower among those infected with Omicron compared to previous variants (15, 19), our results indicate that the burden of Long COVID 90 days after Omicron infection is substantial.

Second, our study extends results from many studies conducted prior to the emergence of Omicron showing the risk of developing Long COVID is higher among females, middle-aged and older adults, and those with pre-existing health conditions, to populations infected by Omicron (13–17).

Third, we found almost 40% of those experiencing Long COVID reported accessing health care for associated ongoing symptoms 60-90 days post diagnosis, a result consistent with a Long COVID study in Switzerland (20). This translates to 1 in every 15 adults diagnosed with COVID-19 in WA during the study period seeking COVID-19-related health care 2-3 months post diagnosis. If this figure is extrapolated to the 1.2 million persons with a first SARS-CoV-2 diagnosis reported in WA in 2022, it equates to approximately 80,000 healthcare encounters. In our setting, the vast majority of health care encounters were visits to a GP, while the impact of Long COVID on ED attendance and hospital admissions was more limited. These data suggest that investments intended to provide ongoing care to persons with Long COVID should include enhanced support for primary care.

Fourth, we found that almost two-thirds of persons with Long COVID who were working or studying at the time of their COVID-19 diagnosis were able to fully return to these activities within a month. Nevertheless, nearly one in five had reduced hours or were not working at 90 days post COVID-19 diagnosis. As reported elsewhere (21), these data highlight that in addition to negative impacts on health, there may be significant economic and workforce implications associated with Long COVID.

Finally, our results add to the growing body of evidence showing that COVID-19 vaccinations are associated with a reduced risk of developing Long COVID; this finding may inform health messaging regarding the benefits of COVID-19 vaccination. (22)

This study has limitations. First, like many other Long COVID investigations, our case definition for ‘Long COVID’ relied on subjective, self-reported, ongoing symptoms and the specificity of these symptoms for diagnosing Long COVID is not well defined. We used WHO’s definition of “post COVID condition” as the basis for our case definition but adapted it by replacing “symptoms lasting for at least 2 months” with a requirement that persons had to be currently experiencing symptoms at the time of the survey to be classified as having Long COVID. This change was made to increase interpretability of the questions posed to participants using a self-administered web-based survey tool but may limit direct comparisons to other studies. Second, because our study population was drawn from persons diagnosed with SARS-CoV-2 who had consented to follow-up for future research, we did not have a control group of respondents without recent SARS-CoV-2 infection; this prevented us from assessing the level of ‘background’ persistent symptomology among those without antecedent COVID-19 illness for comparison. Third, our data on health service utilisation and impacts on work or study were based on self-report and not independently verified by the person’s healthcare provider, employer, or academic institution. Fourth, we did not obtain information on the severity of the initial COVID-19 illness from each participant; but there were only 171 (0.7%) total hospital admissions recorded for the 22,744 persons who were sent the survey SMS; we therefore can infer that the vast majority of the survey respondents were not hospitalised for their illness. Last, persons with chronic illness have been shown to be more likely to be willing to participate in research, and if this occurred in our study it may have inflated the proportion of respondents who met Long COVID case definition (23); however, in our setting the proportion with Long COVID among the ∼80% of respondents with no pre-existing health issues was 16.2%, i.e. a figure closely aligned with the overall estimate of 18.2%

This investigation also has several strengths. First, to minimise selection bias, all adults residing in WA with a notified SARS-CoV-2 infection during the study period who had consented to be contacted for COVID-19 research at the time of their initial illness were eligible for inclusion. In addition, ascertainment of SARS-CoV-2 infections during the study period was likely very high since PCR tests and government-provided free RATs were widely available and subject to mandatory reporting. Second, using resource-efficient communication and data collection technologies, a large cohort of eligible study subjects was able to be sampled, and the impact of potential participation bias was mitigated by achieving a respectable survey response rate (>50%). We also applied sampling weights using age and sex to adjust for over-and under-representation of specific sub-groups. Third, individuals diagnosed with repeat infections were actively excluded and because there was no widespread transmission of SARS-CoV-2 in WA prior to 2022, it is unlikely there were substantial numbers of unrecognised prior infections. Last, to enhance the generalisability of our results, survey questions were taken from previously validated tools where appropriate (3, 8, 24)

In summary, this study demonstrated that in a highly vaccinated population, approximately 1 in 5 persons first infected with the Omicron variant reported symptoms consistent with Long COVID ninety days after their initial COVID-19 diagnosis. Although hospital admissions were uncommon, Long COVID resulted in a substantial burden of GP consultations for ongoing symptoms. The majority of persons with Long COVID reported fully returning to work or study within a month after their illness, but ∼20% reported negative impacts extending beyond three months. Additional follow-up studies will be vital to enhance our understanding of the duration and severity of Long COVID and for identifying effective treatments.

## Supporting information

Appendix 1: Long COVID survey questionnaire

## Data Availability

All data produced in the present work are contained in the manuscript.

## Appendix

**Appendix 2:**
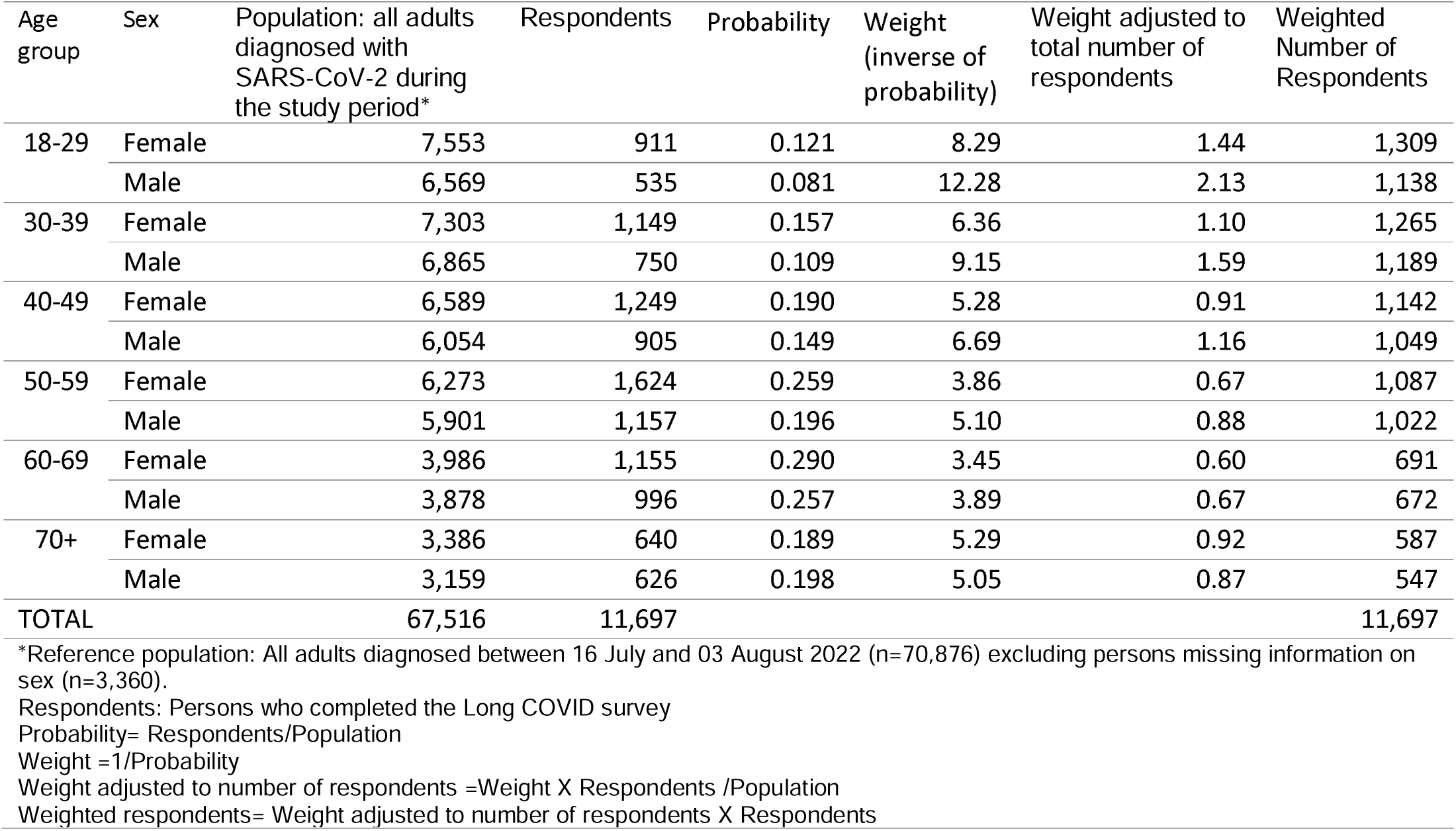
Sampling weight applied to Long COVID online survey in Western Australia, 2022.

| Age group | Sex | Population: all adults diagnosed with SARS-CoV-2 during the study period* | Respondents | Probability | Weight (inverse of probability) | Weight adjusted to total number of respondents | Weighted Number of Respondents |
| --- | --- | --- | --- | --- | --- | --- | --- |
| 18-29 | Female | 7,553 | 911 | 0.121 | 8.29 | 1.44 | 1,309 |
|  | Male | 6,569 | 535 | 0.081 | 12.28 | 2.13 | 1,138 |
| 30-39 | Female | 7,303 | 1,149 | 0.157 | 6.36 | 1.10 | 1,265 |
|  | Male | 6,865 | 750 | 0.109 | 9.15 | 1.59 | 1,189 |
| 40-49 | Female | 6,589 | 1,249 | 0.190 | 5.28 | 0.91 | 1,142 |
|  | Male | 6,054 | 905 | 0.149 | 6.69 | 1.16 | 1,049 |
| 50-59 | Female | 6,273 | 1,624 | 0.259 | 3.86 | 0.67 | 1,087 |
|  | Male | 5,901 | 1,157 | 0.196 | 5.10 | 0.88 | 1,022 |
| 60-69 | Female | 3,986 | 1,155 | 0.290 | 3.45 | 0.60 | 691 |
|  | Male | 3,878 | 996 | 0.257 | 3.89 | 0.67 | 672 |
| 70+ | Female | 3,386 | 640 | 0.189 | 5.29 | 0.92 | 587 |
|  | Male | 3,159 | 626 | 0.198 | 5.05 | 0.87 | 547 |
| TOTAL |  | 67,516 | 11,697 |  |  |  | 11,697 |

\*Reference population: All adults diagnosed between 16 July and 03 August 2022 (n=70,876) excluding persons missing information on sex (n=3,360).
Respondents: Persons who completed the Long COVID survey
Probability= Respondents/Population
Weight =1/Probability
Weight adjusted to number of respondents =Weight X Respondents /Population
Weighted respondents= Weight adjusted to number of respondents X Respondents

