## Appendix 1: Long COVID survey questionnaire for "Long COVID in a highly vaccinated population infected during a SARS-CoV-2 Omicron wave – Australia, 2022"

Do you currently have any new or ongoing symptoms or health problems that you believe are related to your COVID-19 illness?”

 Response options: “Yes”, “No”, “Not sure”

If they select “Yes” or “Not sure”, the survey questions will appear.

“We would like to learn more about the ongoing symptoms or health conditions you may be experiencing due to your COVID illness. (Survey 1.1)

If they select “No”, the following statement will appear.

“We would like to learn about your health status before and after your COVID-19 diagnosis. (Survey 1.2)

**SURVEY 1****.1 (for participants affirming that they have ongoing health issues)**

1) Are you currently experiencing any of the following symptoms?

Tiredness or fatigue that interferes with daily life	☐Yes	☐No	☐Not sure

Headache	☐Yes	☐No	☐Not sure

Fever	☐Yes	☐No	☐Not sure

Difficulty breathing or shortness of breath	☐Yes	☐No	☐Not sure

Cough	☐Yes	☐No	☐Not sure

Chest pain	☐Yes	☐No	☐Not sure

Fast-beating or pounding heart (heart palpitations) ☐Yes	☐No	☐Not sure

Difficulty thinking or concentrating (“brain fog”)	☐Yes	☐No	☐Not sure

Sleep problems	☐Yes	☐No	☐Not sure

Dizziness when you stand up (light-headedness)	☐Yes	☐No	☐Not sure

Memory loss or confusion	☐Yes	☐No	☐Not sure

Pins-and-needles feelings	☐Yes	☐No	☐Not sure

Change in smell or taste	☐Yes	☐No	☐Not sure

Low mood or not enjoying anything	☐Yes	☐No	☐Not sure

Increased worry or anxiety	☐Yes	☐No	☐Not sure

Diarrhea	☐Yes	☐No	☐Not sure

Stomach pain	☐Yes	☐No	☐Not sure

Rash	☐Yes	☐No	☐Not sure

Joint or muscle pain	☐Yes	☐No	☐Not sure

Loss of appetite or eating less than usual	☐Yes	☐No	☐Not sure

Nausea or vomiting	☐Yes	☐No	☐Not sure

Females only - Changes in your menstrual cycle	☐Yes	☐No	☐Not sure

2) Do you think your current symptoms are the result of your recent COVID infection?

☐ Yes – I think they have been caused by my recent COVID infection

☐ No – I think they are likely explained by a separate illness or health condition I have/have had

☐ I am not sure

3) Do you think your current symptoms are consistent with the condition called “Long COVID”?

☐ Yes

☐ No

☐ I am not sure if my symptoms are considered to be ‘Long COVID’

☐ I am not sure what ‘Long COVID’ means

4) In the last month, have you seen a GP because of ongoing symptoms following your COVID illness?

☐ Yes	☐ No	☐ Not sure

5) In the last month, have you been to an emergency department because of ongoing symptoms following your COVID illness?

☐ Yes	☐ No	☐ Not sure

6) In the last month, have you been admitted to hospital because of ongoing symptoms following your COVID illness?

☐ Yes	☐ No	☐ Not sure

7) If you were working and/or studying before your COVID diagnosis, have you been able to continue these activities after your illness?

☐ Yes, I was able to fully return to work/study within a month of my COVID illness

☐ Yes, but I needed more than a month before I was able to fully return to work/studying

☐ Yes, but I have had to reduce the number of hours I work/study

☐ No, I have not returned to work/study

☐ I was not working or studying at the time I developed my COVID illness

☐ I’m not sure

8) How would you describe the outcome of your COVID illness at the present time?

☐ I am fully recovered from my COVID illness now

☐ I am not fully recovered, but I can do my usual activities

☐ I am still recovering, but not able to all my usual activities

☐ I do not feel like I am recovering

☐ I’m not sure

9) How would you describe your overall health status now?
   a. ☐ Excellent
   b. ☐ Good
   c. ☐ Fair
   d. ☐ Poor
   e. ☐ I’m not sure
10) Before you tested positive for COVID-19, did you have any significant or chronic health issues?

☐ Yes	☐ No	☐ Not sure

11) (If yes) Please indicate the type(s) of health issues you had prior to being diagnosed with COVID-19:

☐ Heart problems

☐ Lung problems

☐ Asthma

☐ kidney disease

☐ Gastrointestinal problems such as diarrhea or stomach pain

☐ Diabetes

☐ Obesity

☐ Neurologic problems such as sleep problems, headache, depression, or anxiety

☐ Immune system problems

☐ Cancer

☐ Other

We would like to send you another brief survey three months from today to find out if you have fully recovered from your COVID illness or if you are still having symptoms. Would that be okay with you?

☐ Yes

☐ No

Response to either 🡪 “Thank you for helping the WA Health better understand the longer-term impact of the COVID-19 pandemic. Please click "Finish Survey" below, then click "Next" to submit your answers.

If you have concerns about any ongoing symptoms, or need information on how to manage them, you should speak with your GP or call healthdirect at 1800 022 222.”

END >>>>>>>>>>>>>>>>>>>>>>>>>>>>>>>>>>>>>>>>>>>>>>>>>>>>>>

**SURVEY 1.2 (for participants responding that they DO NOT have ongoing health issues)**

Thank you for taking a few moments to tell us about your current health status.

We would like to learn about your health status before and after your COVID-19 diagnosis. This brief survey will take about 2 minutes.

1) Before you tested positive for COVID-19, did you have any significant or long-standing health issues?

☐ Yes	☐ No	☐ Not sure

2) (If Yes or Not sure) Please indicate the type(s) of health issues you had prior to being diagnosed with COVID-19:
   a. ☐ Heart problems
   b. ☐ Lung problems
   c. ☐Asthma
   d. ☐kidney disease
   e. ☐ Gastrointestinal problems such as Diarrhea or stomach pain
   f. ☐ Diabetes
   g. ☐ Obesity
   h. ☐ Neurologic problems such as sleep problems, headache, depression, or anxiety
   i. ☐ Immune system problems
   j. ☐ Cancer
   k. ☐ Other
3) How would you describe your overall health status now?
   f. ☐ Excellent
   g. ☐ Good
   h. ☐ Fair
   i. ☐ Poor
   j. ☐ I’m not sure
4) If you were working and/or studying before your COVID diagnosis, have you been able to continue these activities after your illness?
   a. ☐ Yes, I was able to fully return to work/study within a month of my COVID illness
   b. ☐ Yes, but I needed more than a month before I was able to fully return to work/studying
   c. ☐ Yes, but I have had to reduce the number of hours I work/study
   d. ☐ No, I have not returned to work/study
   e. ☐ I was not working or studying at the time I developed my COVID illness
   f. ☐ I’m not sure

“Thank you for helping the WA Health better understand the impact of the COVID-19 pandemic.” Please click "Finish Survey" below, the click "Next" to submit your answers.

END >>>>>>>>>>>>>>>>>>>>>>>>>>>>>>>>>>>>>>>>>>>>>>>>>>>>>>
